# “Urban Tertiary Care Centre Experience of Characteristics of Severe COVID-19 Pneumonia”

**DOI:** 10.1101/2021.08.16.21262073

**Authors:** Nehal M Shah, Janakkumar R Khambholja, Nilay Suthar, Hemant Tiwari, Vandit Desai, Vishal Beriwala

**Affiliations:** Department of Medicine, SVP hospital, SMT NHL Municipal Medical College, Ellisbridge, Ahmedabad-380006; SMT NHL Municipal Medical College, Ellisbridge,Ahmedabad; SMT NHL Municipal Medical College, Ellis bridge, Ahmedabad—380006

**Keywords:** Severe COVID-19 Pneumonia, ARDS, Inflammatory Markers, Adjunct Treatment

## Abstract

**Introduction:** The global pandemic of novel coronavirus disease 2019 (COVID-19) caused by severe acute respiratory syndrome coronavirus 2 (SARS-CoV-2) began in Wuhan, China, in December 2019, and has since spread worldwide.^[1]^ This study attempts to summarize current evidence regarding major inflammatory markers, severity predictors and its impact on outcome, which provide current clinical experience and treatment guidance for this novel coronavirus.

**Methods:** This is a retrospective observational study done at an urban teaching covid-19 designated hospital. Hospital data were analysed with aim of studying inflammatory markers, predictors and outcome. Patients were classified in Mild, Moderate, Severe & Critical categories of COVID cases. Their clinical parameters, laboratory investigations, radiological findings & Outcome measures were studied. Strength of association & correlation of those parameters with severity and in-hospital mortality were studied.

**Results:** A total 204 (N) patients were clinically classified into different severity groups, as per MOHFW and qCSI(quick Covid Severity Index) guidelines, as Mild (34), Moderate (56), Severe (39) and Critical (75). The mean(SD) age of the cohort was 55.1+13.2 years; 74.02% were male. Severe COVID-19 illness is seen more in patients more than 50 years of age. COVID-19 patients having IHD develop worse disease with excess early in-hospital mortality. Respiratory rate & Heart Rate on admission are correlated with severe and stormy disease. Among Inflammatory markers, on admission LDH, D-Dimer and CRP are related with severity and excess in-hospital death rate.

**Conclusion:** Advanced age, male gender, IHD, Respiratory Rate & Heart Rate on admission were associated with severe covid-19 illness. S. Lactate Dehydrogenase & D-dimer was associated with severe covid-19 illness and early in-hospital death.

## INTRODUCTION

The global pandemic of novel coronavirus disease 2019 (COVID-19) caused by severe acute respiratory syndrome coronavirus 2 (SARS-CoV-2) began in Wuhan, China, in December 2019, and has since spread worldwide.^[1,2]^ The viral genome of SARSCoV-2 was rapidly sequenced to enable diagnostic testing, epidemiologic tracking, and development of preventive and therapeutic strategies.^[3]^This study attempts to summarize current evidence regarding major inflammatory markers, severity predictors clinical experience and treatment guidance for this pandemic coronavirus.

## METHODS

This is a retrospective observational study conducted at a tertiary care Ahmedabad Municipal Corporation run COVID-19 designated hospital. Cases were enrolled from Hospital Computerised Data Management System from June 2020 to August 2020. Cases with Different severity of COVID-19 patients were classified according to institutional SOP, as per guidance from MOHFW^[4]^.

| CRITERIA | MILD | MODERATE | SEVERE | CRITICAL |
| --- | --- | --- | --- | --- |
| RR/ O<sub>2</sub>Support | Normal/ No | 24-30/ Nasal prongs | ≥30/ Mask | HFNC/NIV/IMV |
| SpO<sub>2</sub> | >94 | 90-94 | <90 | <90 |
| O<sub>2</sub> requirement | No | 2-4 L/min | ≤10 L/min | >10 L/min |

Case definitions:<sup>[4,5]</sup>

Patients were studied for baseline characteristics, laboratory and radiological characteristics. This study attempts to summarize current evidence regarding major inflammatory markers, severity predictors and its impact on outcome, which provide current clinical experience and treatment guidance for this novel coronavirus. Study looked at outcomes as Death or Discharge and studied its association with several possible predictors. In-hospital mortality was defined as death during hospitalisation for covid-19 illness. Correlation with severity and in-hospital death were also accessed.

### Statistical analysis

Looking to approximately twenty per cent of in-hospital mortality rate in covid-19 cases^[6,7]^, 80% of Power of study & Two sided alpha error of 5%, required sample size is 193. Keeping in view several possibilities we kept effective sample size of 204 for the current study. Normality of the distribution was tested and Kurtosis was calculated. Categorical variables are reported as number (%). Normally distributed continuous data were reported as mean ± standard deviation (SD).Nonparametric data were analysed by Chi square method. Parametric data were analysed by standard students t test and ANOVA. Pearson Correlation was used wherever applicable. Binary Logistic Regression was applied for predicting dichotomous binary dependant outcomes. Receiver operating characteristic(ROC) curves were used to evaluate the potential predictive value of inflammatory markers on prognosis of hospitalised COVID-19 patients. The Hosmere-Lemeshow test was used to calibrate the ROC curves. Statistical software were used as per necessity like SPSS v25/v26, GraphPad Prism, MedCalc& MS Excel 365. *P*-values less than 0.05 were considered as statistically significant difference.

## RESULTS

This is an observational study conducted at a tertiary care urban teaching hospital. The study population characteristics and number can be inferred from Figure-1.

**Figure-1.**
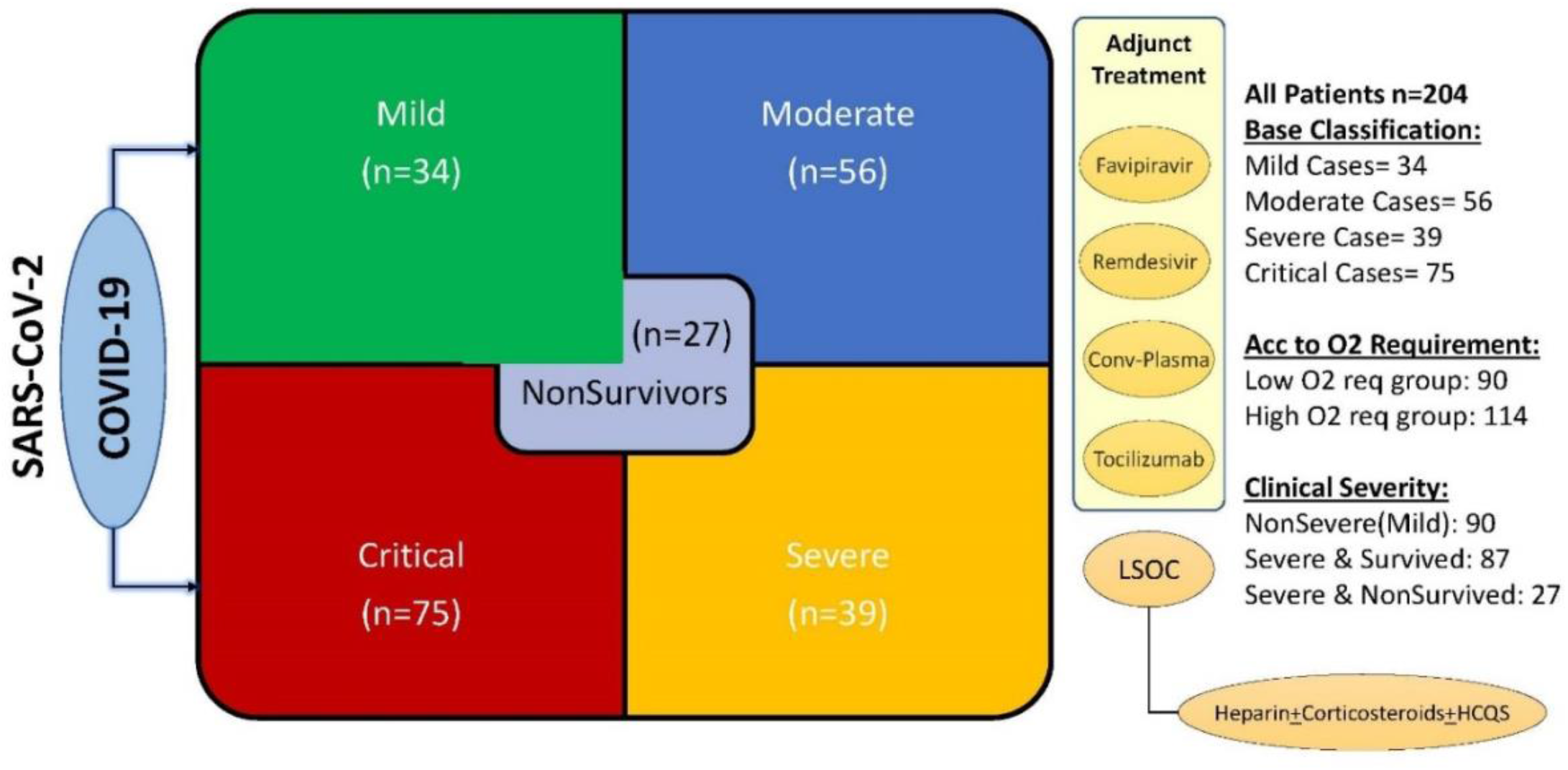
Patient groups and Clinical Categories

### Baseline characteristics

#### 1. Age and Gender

Study period was done from June 2020 to August 2020. All the data of vitals, radiological and pathological tests, clinical notes and relevant treatment records of hospitalized confirmed COVID-19 adult patients were documented to iHIS, which were transferred to Google forms for data collection. Data entry and cross verification of each record were done to minimise human errors. A total 204 (N) patients were studied, and they were clinically classified into different severity groups, as per MOHFW and qCSI (quick Covid Severity Index) guidelines, as Mild (34), Moderate (56), Severe (39) and Critical (75), The mean(SD) age of the cohort was 55.1+13.2 years; 74.02% were male. When we considered age group more than 50years, it comprises maximum Critical (37%) cases followed by Moderate (17%) illness compared to patients aged less than 50 years of age. 65 % of cases were aged more than 50 years of age (*P*=0.024).

Furthermore, these four clinical categories ie: mild, moderate, severe and critical were clubbed to sub-groups. ‘NonSevere’ cases are in Gr-A,’Severe & Survived’cases are in Gr-B and ‘Severe & NonSurvived’ are in Gr-C (Table-1.1) When considering 50years as the cut-off, patients more than 50 years had higher pulmonary compromise in the form of higher need of oxygen or assistive mechanical ventilator support.(*P*<0.001)

**Table-1.** Baseline Demographic Details of Study population

|  |  | All | Mild | Moderate | Severe | Critical |
| --- | --- | --- | --- | --- | --- | --- |
|  |  | n (%)<br>mean±SD | n (%)<br>mean±SD | n (%)<br>mean±SD | n (%)<br>mean±SD | n (%)<br>mean±SD |
| Age-Groups | 0 to 49 years | 69 (33.82%) | 12 (35.29%) | 16 (28.57%) | 16 (41.03%) | 25 (33.33%) |
|  | 50 to 64 years | 85 (41.67%) | 12 (35.29%) | 24 (42.86%) | 14 (35.90%) | 35 (46.67%) |
|  | >64 years | 50 (24.51%) | 10 (29.41%) | 16 (28.57%) | 9 (23.08%) | 15 (20.00%) |
|  | All | 204 (100%) | 34 (100%) | 56 (100.00%) | 39 (100.00%) | 75 (100.00%) |
| Gender | Female | 53 (25.98%) | 8 (23.53%) | 12 (21.43%) | 12 (30.77%) | 21 (28.00%) |
|  | Male | 151 (74.02%) | 26 (76.47%) | 44 (78.57%) | 27 (69.23%) | 54 (72.00%) |
|  | All | 204 (100%) | 34 (16.67%) | 56 (27.45%) | 39 (19.12%) | 75 (36.76%) |
| DM |  | 109 (53.43%) | 12 (11.01%) | 35 (32.11%) | 25 (22.94%) | 37 (33.94%) |
| HTN |  | 106 (51.96%) | 12 (11.32%) | 29 (27.36%) | 24 (22.64%) | 41 (38.68%) |
| IHD |  | 27 (13.24%) | 2 (7.41%) | 4 (14.81%) | 5 (18.52%) | 16 (59.26%) |
| HR (on admission) |  | 94.84±15.1 | 94.21±12.6 | 93.63±14.0 | 91.18±13.4 | 97.95±17.2 |
| SBP (on admission) |  | 127.24±14.3 | 125.21±11.9 | 127.75±12.2 | 126.38±16.4 | 128.23±15.6 |
| DBP (on admission) |  | 79.89±7.8 | 80.24±5.0 | 80.29±7.5 | 78.82±7.6 | 80.00±9.1 |
| RR (on admission) |  | 19.68±6.0 | 16.44±1.3 | 18.95±4.1 | 20.38±5.6 | 21.33±7.8 |
| SpO2 (on admission) |  | 92.76±9.0 | 97.03±1.7 | 94.77±4.1 | 91.15±9.5 | 90.16±11.9 |

**Table-1.1.**
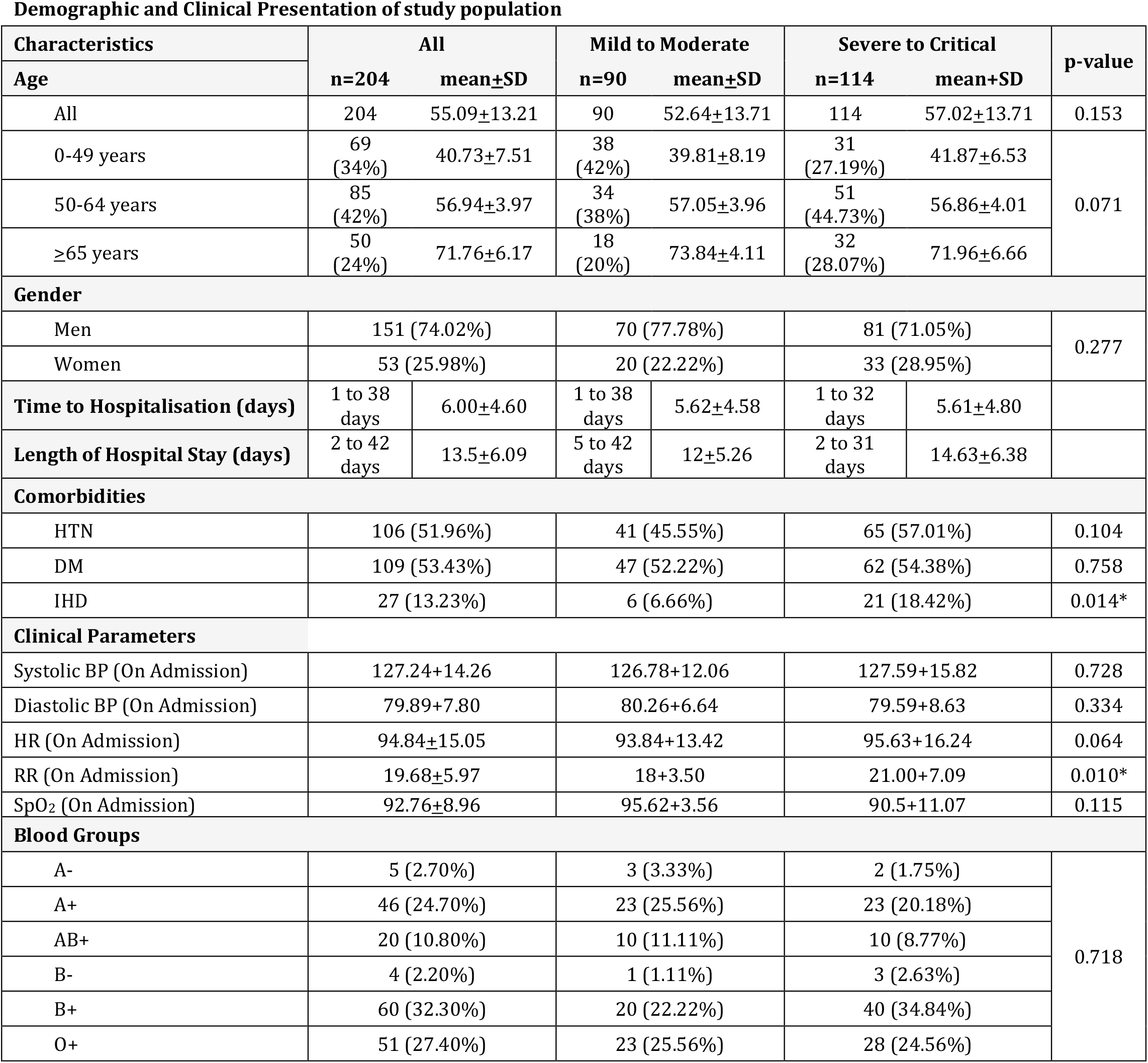
Demographic Details and Baseline Clinical Characteristics

Table-1 Baseline Demographic Details of Study population
| Demographic and Clinical Presentation of study population |  |  |  |  |  |  |  |
| --- | --- | --- | --- | --- | --- | --- | --- |
| Characteristics | All |  | Mild to Moderate |  | Severe to Critical |  | p-value |
| Age | n=204 | mean±SD | n=90 | mean±SD | n=114 | mean+SD |  |
| All | 204 | 55.09±13.21 | 90 | 52.64±13.71 | 114 | 57.02±13.71 | 0.153 |
| 0-49 years | 69<br>(34%) | 40.73±7.51 | 38<br>(42%) | 39.81±8.19 | 31<br>(27.19%) | 41.87±6.53 | 0.071 |
| 50-64 years | 85<br>(42%) | 56.94±3.97 | 34<br>(38%) | 57.05±3.96 | 51<br>(44.73%) | 56.86±4.01 |  |
| ≥65 years | 50<br>(24%) | 71.76±6.17 | 18<br>(20%) | 73.84±4.11 | 32<br>(28.07%) | 71.96±6.66 |  |
| Gender |  |  |  |  |  |  |  |
| Men | 151 (74.02%) |  | 70 (77.78%) |  | 81 (71.05%) |  | 0.277 |
| Women | 53 (25.98%) |  | 20 (22.22%) |  | 33 (28.95%) |  |  |
| Time to Hospitalisation (days) | 1 to 38<br>days | 6.00±4.60 | 1 to 38<br>days | 5.62±4.58 | 1 to 32<br>days | 5.61±4.80 |  |
| Length of Hospital Stay (days) | 2 to 42<br>days | 13.5±6.09 | 5 to 42<br>days | 12±5.26 | 2 to 31<br>days | 14.63±6.38 |  |
| Comorbidities |  |  |  |  |  |  |  |
| HTN | 106 (51.96%) |  | 41 (45.55%) |  | 65 (57.01%) |  | 0.104 |
| DM | 109 (53.43%) |  | 47 (52.22%) |  | 62 (54.38%) |  | 0.758 |
| IHD | 27 (13.23%) |  | 6 (6.66%) |  | 21 (18.42%) |  | 0.014* |
| Clinical Parameters |  |  |  |  |  |  |  |
| Systolic BP (On Admission) | 127.24±14.26 |  | 126.78±12.06 |  | 127.59±15.82 |  | 0.728 |
| Diastolic BP (On Admission) | 79.89±7.80 |  | 80.26±6.64 |  | 79.59±8.63 |  | 0.334 |
| HR (On Admission) | 94.84±15.05 |  | 93.84±13.42 |  | 95.63±16.24 |  | 0.064 |
| RR (On Admission) | 19.68±5.97 |  | 18±3.50 |  | 21.00±7.09 |  | 0.010* |
| SpO2 (On Admission) | 92.76±8.96 |  | 95.62±3.56 |  | 90.5±11.07 |  | 0.115 |
| Blood Groups |  |  |  |  |  |  |  |
| A- | 5 (2.70%) |  | 3 (3.33%) |  | 2 (1.75%) |  | 0.718 |
| A+ | 46 (24.70%) |  | 23 (25.56%) |  | 23 (20.18%) |  |  |
| AB+ | 20 (10.80%) |  | 10 (11.11%) |  | 10 (8.77%) |  |  |
| B- | 4 (2.20%) |  | 1 (1.11%) |  | 3 (2.63%) |  |  |
| B+ | 60 (32.30%) |  | 20 (22.22%) |  | 40 (34.84%) |  |  |
| O+ | 51 (27.40%) |  | 23 (25.56%) |  | 28 (24.56%) |  |  |

**Table-1.2.**
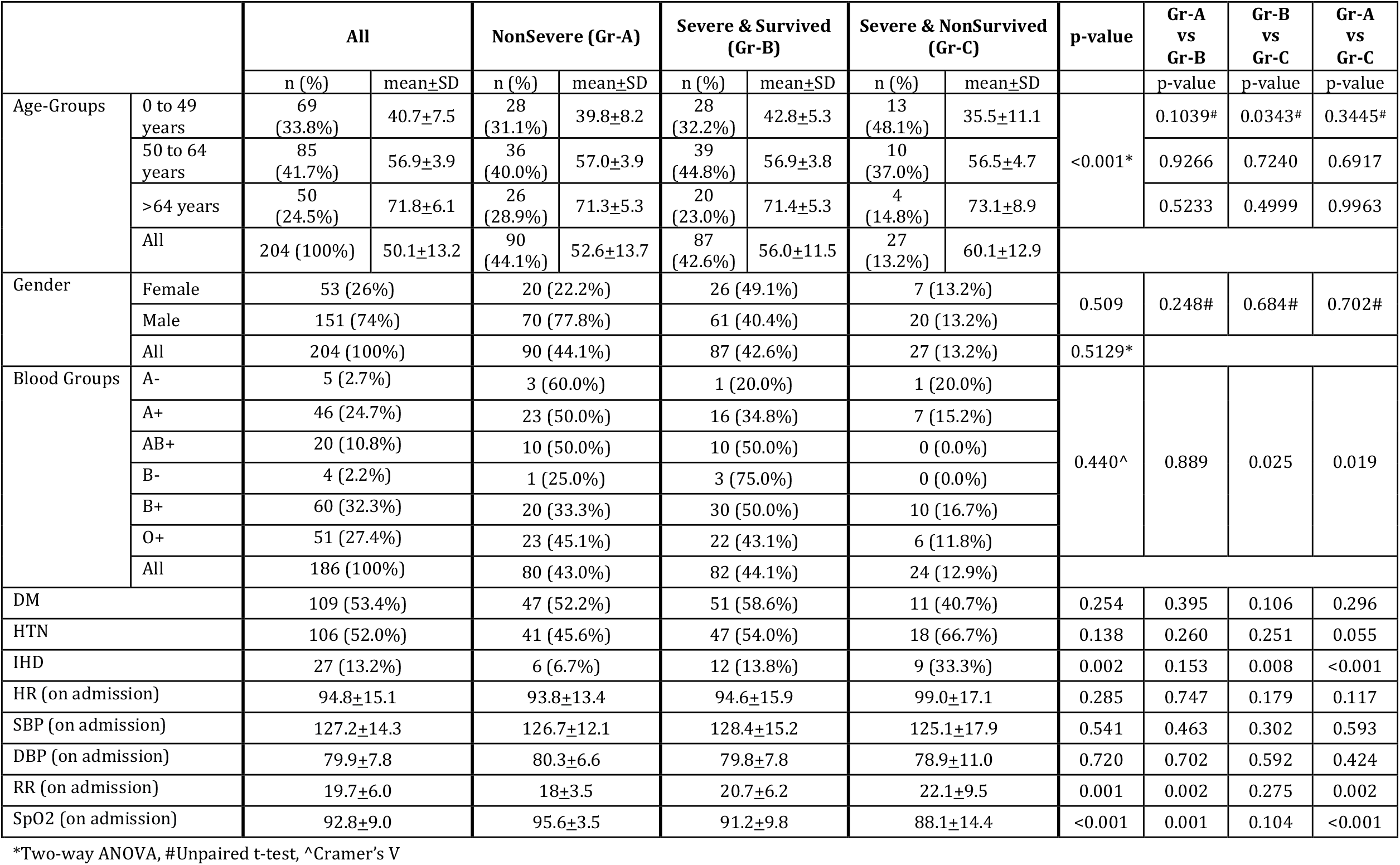
Clinical Classification of COVID Patients in the Study

#### 2. Co-morbidities, vitals, blood groups & COVID Illness

Gr-B and Gr-C cases had more Hypertension (HTN) than Gr-A. Type II diabetes (DM) did not have significant difference between both groups. However, *P*-value (0.0140) was observed to be statistically significant among Ischemic Heart Disease patients’ groups.

Among the vitals on admission of these patients, we found that the difference between the mean Respiratory Rate (*P*-value= 0.0100) was statically significant between both groups but Heart Rate (*P*-value=0.0644) was not. Blood Group analysis of these patients showed that highest cases of covid-19 pneumonia had B Positive blood group followed by O positive, A positive and AB positive.

### NonSevere (Gr-A), Severe & Survived (Gr-B) and Severe &NonSurvived (Gr-C) Sub-Group Patients’ analysis

The overall age difference in the mean age between the groups (Gr-A, Gr-B &Gr-C) was found to be highly statistically significant (*P*<0.001) in our study population. In between these three groups, we compared the mean difference of age among different age groups, but only the mean difference was statistically significant, in the age group of less than 50years, between Gr-B and Gr-C. Population aged more than 50 years of age have higher chances of having severe form of COVID-19 pneumonia [n=83(61%)] compared to Gr-A disease [n=52(39%)] which is statistically significant (*P*=0.023).. But, among patients aged >50years, male gender has a higher chance of developing severe form of covid-19 illness compared to female gender [59(44%) vs 24(18%)]. Although it was statistically not significant (*P*=0.619).

When we studied correlation of blood groups with the severity, it was observed in current study population that it was statistically highly significant between Gr-A vs Gr-C (*P*=0.019) and Gr-B vs Gr-C (*P*=0.025).

Among Gr-C patients, the commonest comorbidity observed is Hypertension (HTN) (67%) followed by Diabetes Mellitus (DM) (41%) and then Ischemic Heart Diseases (IHD) (33%). Although, the findings were statistically significantly associated with clinical severity only in case of IHD (*P*=0.002). On Admission Respiratory Rate (RR) and SpO_2_ were found statistically significant as overall clinical severity of covid-19 cases and among Gr-A vs Gr-B and Gr-B vs Gr-C. Rest of other observed clinical signs were not found to have significance

### Measured Laboratory Parameters& Chest X-rays

We observed several laboratory parameters at the time of hospital admission and during hospital course. Table-2 describes the detailed observations in various clinical categories. The mean Haemoglobin, Platelet count & total white blood cell count were not much different in all the categories on admission. The NLR (Neutrophil-Lymphocyte Ratio) was higher among the severe and critical categories of cases. All other observed laboratory parameter were found statistically not significant among these four categories.

**Table-2.** Disease Clinical Categories and Measured Laboratory Parameters

Table-1.2 Clinical Classification of COVID Patients in the Study
|  |  | Disease Clinical Categories |  |  |  |  |  |  |  | p-value |
| --- | --- | --- | --- | --- | --- | --- | --- | --- | --- | --- |
|  |  | Mild |  | Moderate |  | Severe |  | Critical |  |  |
|  |  | Mean±SD | N(%) | Mean±SD | N(%) | Mean±SD | N(%) | Mean±SD | N(%) |  |
| Hb on Admission |  | 13±1 |  | 13±2 |  | 12±2 |  | 13±2 |  | 0.294 (NS) |
| TC on Admission |  | 5438±2058 |  | 7166±3168 |  | 7352±4236 |  | 8025±3952 |  | 0.288 (NS) |
| On admission NLR |  | 5.15±3.88 |  | 6.96±7.22 |  | 7.93±5.43 |  | 7.81±6.86 |  | 0.350 (NS) |
| Platelet Count (Lakh) |  | 242764±70568 |  | 252732±110253 |  | 269666±130071 |  | 249466±99587 |  | 0.703 (NS) |
| IL-6 on Admission |  | 22±20 | 34(17%)* | 24±22 | 56(27%)* | 34±39 | 39(19%)* | 101±177 | 75(37%)* | 0.570 (NS) |
| NT Pro BNP |  | 211±224 | 34(17%)* | 623±854 | 56(27%)* | 484±858 | 39(19%)* | 1659±2371 | 75(37%)* | 0.467 (NS) |
| Blood Groups | A- |  | 2(6.7%)# |  | 1(2.0%)# |  | 0(0.0%)# |  | 2(2.9%)# | 0.718 (NS) |
|  | A+ |  | 9(30.0%)# |  | 14(28.0%)# |  | 7(19.4%)# |  | 16(22.9%)# |  |
|  | AB+ |  | 4(13.3%)# |  | 6(12.0%)# |  | 5(13.9%)# |  | 5(7.1%)# |  |
|  | B- |  | 0(0.0%)# |  | 1(2.0%)# |  | 0(0.0%)# |  | 3(4.3%)# |  |
|  | B+ |  | 9(30.0%)# |  | 11(22.0%)# |  | 16(44.4%)# |  | 24(34.3%)# |  |
|  | O+ |  | 6(20.0%)# |  | 17(34.0%)# |  | 8(22.2%)# |  | 20(28.6%)# |  |
| Chest X-ray findings* |  |  |  |  |  |  |  |  |  |  |
| B/L GGO |  | 6(2.9%) |  | 15(7.4%) |  | 11(5.4%) |  | 13(6.4%) |  | 0.166 (NS) |
| B/L GGO + Interstitial thickening |  | 3(1.5%) |  | 1(0.5%) |  | 1(0.5%) |  | 1(0.5%) |  |  |
| B/L whiteout lungs |  | 3(1.5%) |  | 1(0.5%) |  | 0(0.0%) |  | 6(2.9%) |  |  |
| GGO + Patchy consolidation |  | 21(10.3%) |  | 36(17.6%) |  | 27(13.2%) |  | 54(26.5%) |  |  |
| Normal |  | 1(0.5%) |  | 3(1.5%) |  | 0(0.0%) |  | 1(0.5%) |  |  |

Chest X-rays of Severe and Critical COVID-19 patients had findings of bilateral ground glass opacities with patchy consolidations; 26% in Critical covid-19 cases and 13% in severe covid-19 cases. Although this finding was found to be statistically not significant.

### Inflammatory markers and Clinical Severity in COVID-19 Pneumonia

Figure-2 depicts the trend of various inflammatory markers and its association with clinical severity (Gr-A, Gr-Band Gr-C) over one week since admission. It has been observed that overall CRP remained elevated in Gr-C COVID 19 pneumonia patients in comparison with Gr-A & Gr-B patients (Figure-2). It was observed that in reference to Gr-C patients, CRP-D7 is significantly high compared to Gr-A patients (OR= 1.03, CI: 1.004-1.067, *P*=0.027). On studying the statistical correlation of CRP values with Gr-C category of COVID-19 illness, 89.3% of variance (R^2^=0.893) can be explained by variance in CRP.

**Figure-2.**
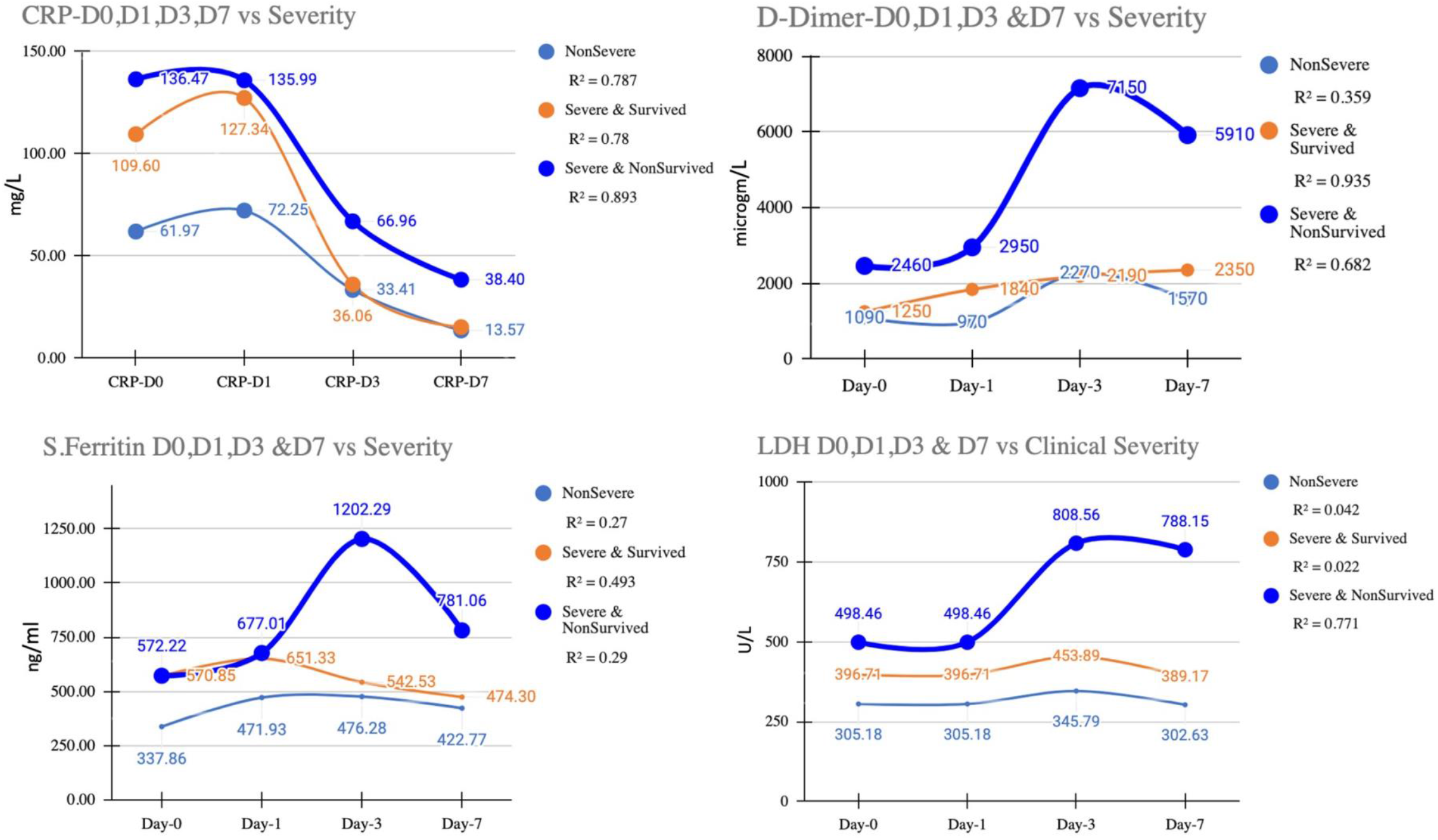
Inflammatory markers and Clinical Severity in COVID-19 Pneumonia

In reference to D-Dimer values, D-Dimer remained elevated in Gr-C COVID 19 pneumonia patients in comparison with Gr-A and Gr-B patients. It was observed that in reference to Gr-A patients, D-Dimer-D7 is statistically significant in Gr-C patients (OR= 1.42, CI: 1.100-1.836, *P*=0.007). On studying the statistical correlation of D-Dimer values with Gr-C category of COVID-19 illness, 68.2% of variance (R^2^=0.682) can be explained by variance in D-Dimer values.

In reference to S. Ferritin values, S. Ferritin remains elevated in Gr-C patients in comparison with Gr-A and Gr-B patients. On studying the statistical correlation of S. Ferritin values with Gr-C category of COVID-19 illness, only 29% of variance (R^2^=0.290) can be explained by variance in S. Ferritin values. When values of S. Ferritin plotted with dichotomous outcomes (Death or Discharge) 88.5% variance (R^2^= 0.885) in outcome would have explained by S. Ferritin values over Day-0 to Day-7, i.e. First week of COVID-19 hospitalization.

In reference to LDH values, LDH remains elevated in Gr-C patients. It was observed that in reference to Gr-A patients, LDH -D7 and LDH-D1 was associated statistically significantly with Gr-C patients with OR= 1.01, CI: 1.008-1.024, p<0.001 and OR= 1.01, CI: 1.003-1.020, *P*=0.007 respectively. On studying the statistical correlation of LDH values with Gr-C category of COVID-19 illness, 77.1% of variance in Severity (R^2^=0.771) could be explained by variance in LDH values. When comparing these four common inflammatory markers with dichotomous outcome (Death vs Discharge), it has been observed that trend of LDH and S.CRP values found to explain more than 85% variance observed in the patients outcome. (LDH R^2^=0.856, S.CRP R^2^=0.893)

### Receiver Operating Characteristic Curve (ROC) and Area under the ROC Curve (AUC)

For better interpretation of these markers, it’s possible utility in predicting the severity of disease (Figure: 3) and outcome (Figure: 4) of covid-19 patients, we plotted ROC curve. We calculated AUC and Cut off value for aforesaid markers. Here, we plotted ROC according to the Day of the measurement of the Parameters (ie: Day-0, Day-1, Day-3 and Day-7).

**Figure-3.**
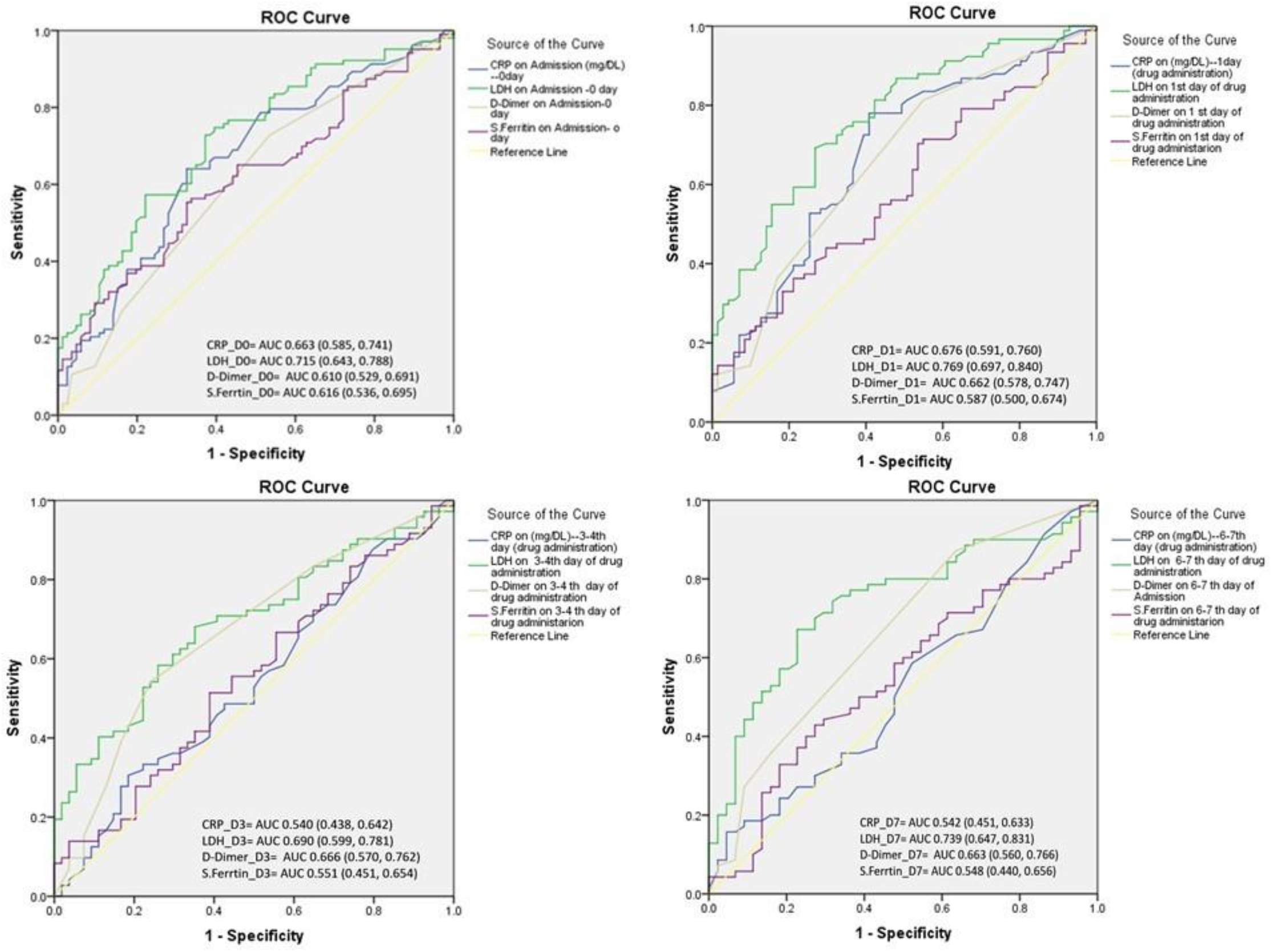
ROC Curve and AUC of Inflammatory Markers vs Severe COVID-19 Pneumonia

**Figure-4.**
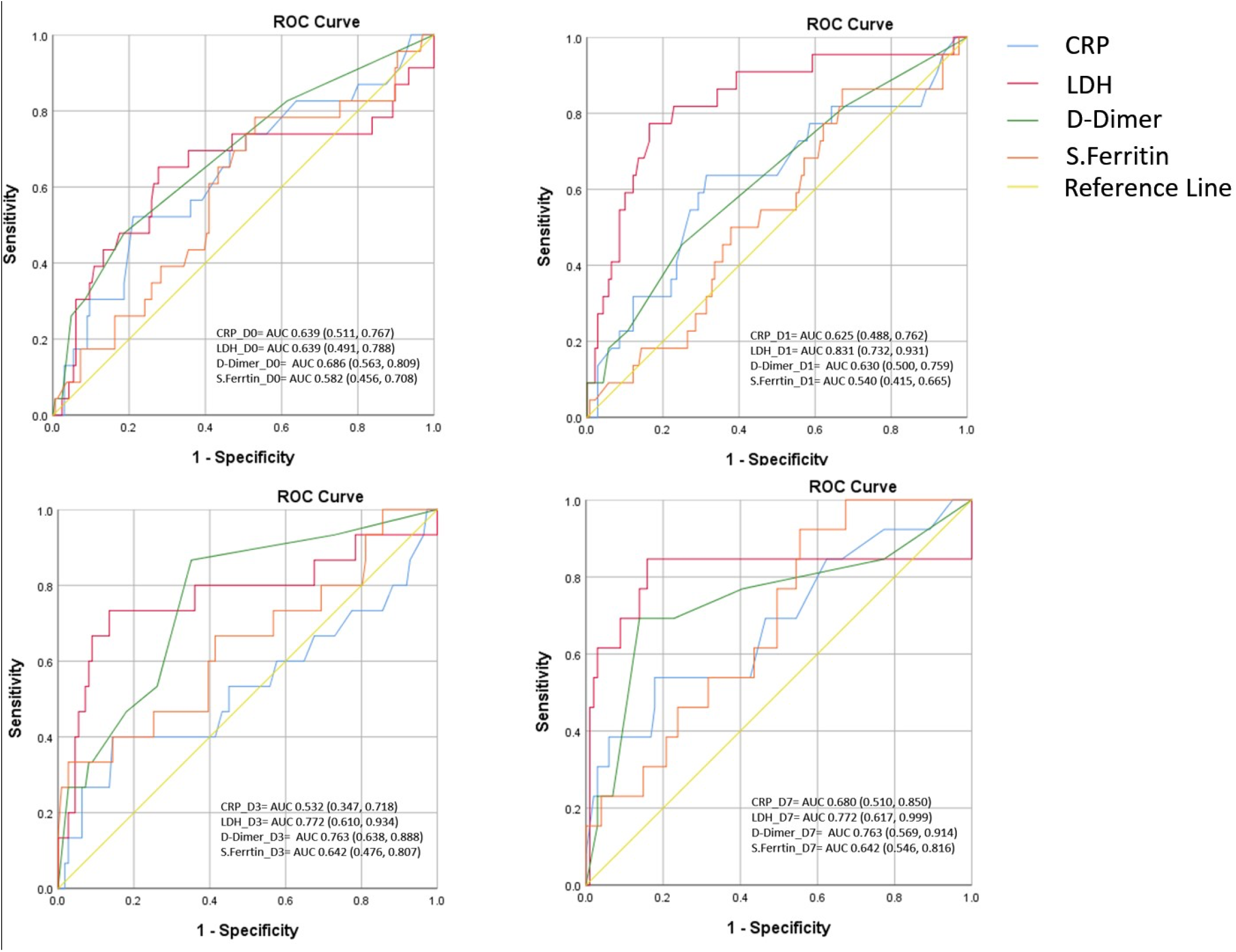
ROC Curve and AUC of Inflammatory Markers vs Adverse Outcome (Death)

Among all inflammatory markers, LDH on all days has shown fair AUC values associated with severity ranging from 0.690 to 0.769. When these markers were used to find AUC of ROC for identifying patients with or without Adverse Outcome (Death) in COVID-19 pneumonia, LDH and D-Dimer were found useful (Figure-4). AUC values for LDH were associated with adverse outcome are ranging from 0.639 to 0.831 while D-Dimer values were ranging from 0.630 to 0.763.

CRP and S. Ferritin were also analysed but their AUC, C-index were not found statistically significant in correlation with severity and outcome.

As per multivariate analysis of mortality and its association with risk factors-age≥50 years (OR=3.34,95% CI 1.105: 10.074, *P*=0.032), Ischemic heart disease (OR=4.417, 95% CI 1.730: 11.272, *P*=0.002) and raised Troponin I, a cardiac injury bio-marker(OR=29.382, 95% CI 8.870: 97.326, *P*<0.001) were significantly associated with excess mortality in COVID -19 pneumonia patients. Male gender, hypertension, Diabetes Mellitus, Vitamin D deficiency (<30 ng/ml), raised D-Dimer (>1000microg/L) and raised uric acid(>6.5 mg/dl) have a trend of excess mortality but they were not found to be statistically significant (Figure-5).

**Figure-5.**
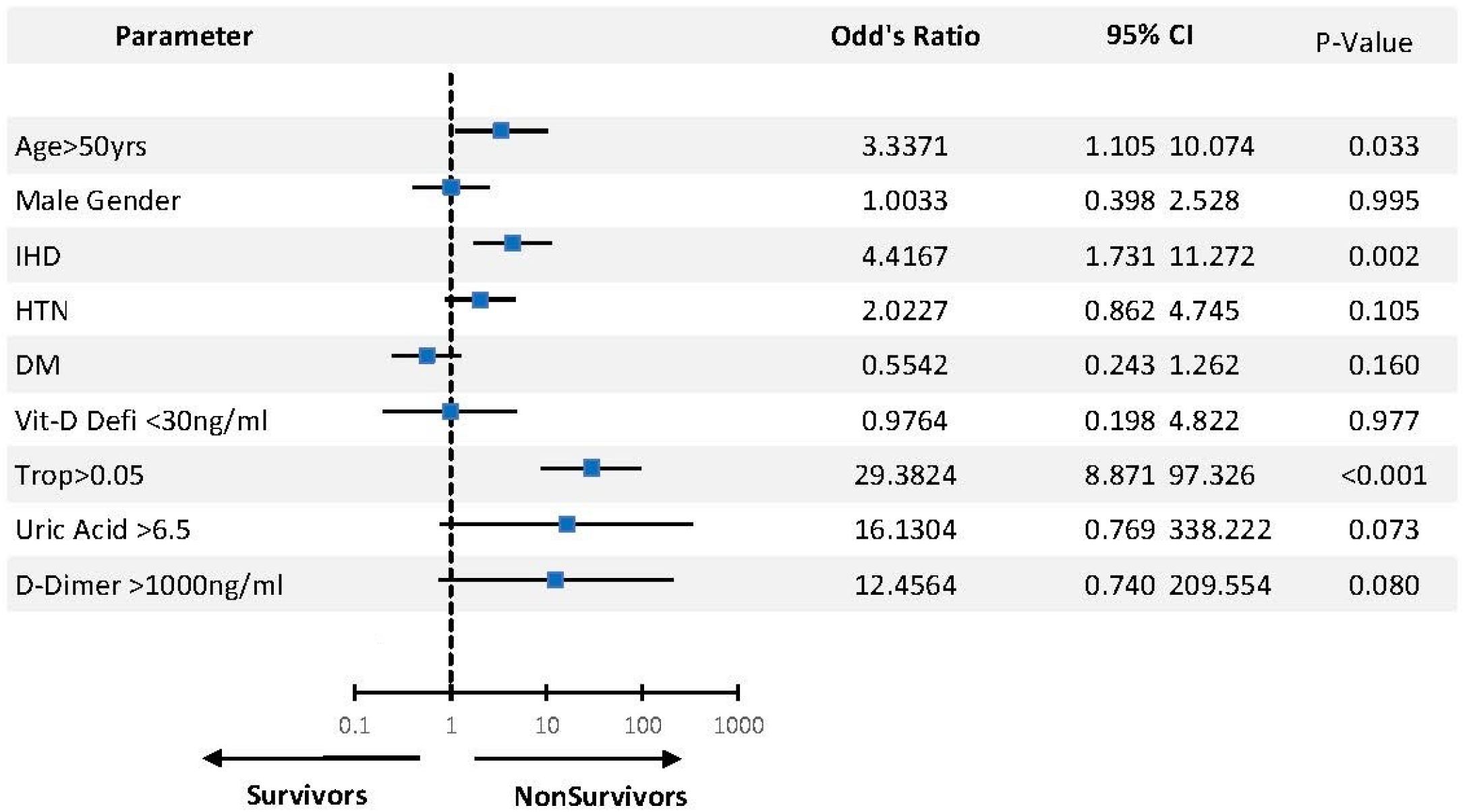
Multivariate analysis of Mortality risk factors for patients with COVID-19 pneumonia

### Predictors and its relationship with Severe COVID-19 Diseases & In-Hospital Death-Mortality in COVID-19 Cases (Logistic Regression Analysis)

Binary Logistic Regression analysis was conducted in SPSS v25 & GraphPad Prism v9. The results were statistically significant in predicting severity and death (Table-3). Significant regression model (p<0.0001) was observed for predicating In-hospital Death (Table-3). This model included LDH-D0, LDH-D1 and LDH-D7 during indoor care of the patients. The pseudo R^2^ (Nagelkerke R Square) of the model is 0.578. The final logistic regression predictive model formula found is, In-hospital Death= Intercept(β0) + (β1)LDH-D0 + (β2)LDH-D1 + (β3)LDH-D7 ie: In-hospital Death= (−7.333) + (−0.01305)LDH-D0 + (0.009151)LDH-D1 + (0.01235)LDH-D7. Survival was coded as 0 and death was coded as 1. The Predicting model shown AUC (C-Index) of 0.9290 (SE: 0.05023, CI: 0.8306 to 1.000, p<0.0001) with Negative Predictive Power of 96% and Positive Predictive Power of 70%.

**Table-3.** Bivariate Multiple Logistic regression analysis predicting In-hospital Death & Severe COVID-19 Disease

| Bivariate Multiple Logistic Regression analysis predicting In-hospital Death |  |  |  |  |  |
| --- | --- | --- | --- | --- | --- |
| | $\beta$ | S.E. | Sig. | Exp($\beta$) | 95% CI |
| LDH-D0 | -0.0130 | 0.0040 | 0.0014 | 0.9870 | 0.977 to 0.994 |
| LDH-D1 | 0.0091 | 0.0033 | 0.0058 | 1.0090 | 1.004 to 1.018 |
| LDH-D7 | 0.0123 | 0.0031 | <0.0001 | 1.0120 | 1.007 to 1.020 |
| Constant | -7.3330 | 1.6760 | <0.0001 | 0.0006 | 1.275e-005 to 0.011 |
| Bivariate Multiple Logistic Regression analysis predicting Severe COVID-19 Disease |  |  |  |  |  |
| | $\beta$ | S.E. | Sig. | Exp($\beta$) | 95% CI |
| LDH_D1 | 0.0044 | 0.0018 | 0.0170 | 1.0040 | 1.001 to 1.008 |
| LDH_D7 | 0.0056 | 0.0018 | 0.0025 | 1.0060 | 1.002 to 1.010 |
| Constant | -3.3800 | 0.9595 | 0.0004 | 0.0340 | 0.004 to 0.194 |

Regarding predicting severity, final logistic regression model formula found : Severe COVID-19 Cases= Intercept(β0) + (β1)LDH_D1 + (β2)LDH_D7 ie: Severe COVID-19 Cases= (−3.380)+(0.004438)xLDH-D1+(0.005640)LDH-D7.Nonsevere category was coded as 0 and severe category was coded as 1. The Predicting model shown AUC (C-Index) of 0.7433(SE: 0.04526, CI: 0.6546 to 0.8320, p<0.0001) with Negative Predictive value of 67% and Positive Predictive Value of 71%.

Hence, it can be inferred that LDH is one of the major markers which can help a clinician to predict severity and adverse outcome in severe covid-19 cases, than any other markers.

## DISCUSSION

Since the origin of SARS-CoV-2 caused COVID-19 infection, stakeholders alerted the whole world about the beginning of a new pandemic. Dearth of research papers flourished the scientific world publication media. A lot of information helped improved management of patients. But regionally relevant research always found lacking in strength of evidence. We attempted to fulfil some gap in these concern and tried to improve patient care.

### Baseline characteristics

In our study, all mild cases (Nonsevere) were presented with mild symptoms and had good prognosis, they mostly recovered within 10 days of symptoms onset and discharged uneventfully. Rapid clinical deterioration was observed in severe cases in second week of illness onset, preceded with rise in inflammatory markers. So thorough risk assessment in early stage of disease might help in predicting disease severity.^[8]^

In our study, patients aged more than 50 years (n-135), 73(54.07%) having severe covid-19 illness (Gr-B+ Gr-C) and fatality rate was 14 (19.18%) among them. Which is almost 10% of all cases above 50 years. Death rates correlated with advanced age as found in other published studies. Overall case fatality rate was 13% in our study in patients aged over 50 years in having severe covid-19 disease. This rate was found higher in several other studies done in China.^[9,10,11,12]^

Gender difference in form of male predominance was seen in our study but fatality rate is almost equal in both genders. Reason for higher male predominance in the disease could be due to enhanced action of transmembrane protease serine 2 (TMPRSS2), which cleaves angiotensin converting enzyme-2 (ACE2) receptors and prime spike proteins.^[13]^ Global data shows higher COVID 19 case fatality rate in male ranging from 1.0 to 3.5 in high income countries while in our country CFR is higher in women due to less detection ratio and delayed hospitalization.^[9,14]^

In our study, symptomatic patients with Rh positive blood types were more likely to be tested positive with COVID-19 compared to Rh negative type.^[15]^But it has no correlation with severity of COVID-19 illness. Studies in Italy and Spain demonstrated higher respiratory failure in “A+ve” blood group, while lower risk was found in “O+ve” blood group patients.^[16]^ In ours study population, “B+ve” blood group was found to have higher number of deaths in comparison to other blood groups, although this needs further studies.

Most common comorbidity identified were Diabetes Mellitus 109(53.43%) followed by Hypertension 106(51.96%) and Ischemic Heart Diseases 27(13.23%). Comorbidities were almost similar in study by Adekunle et al.^[17]^Those who have more than one comorbidities, mortality is high. Similar findings were also observed in a nationwide study conducted at hospitals throughout China and other studies.^[18,19]^ The odds ratio for Ischemic Heart Disease was suggestive of poor outcome in COVID-19 cases.Tachypnoea on admission is associated with poor outcome. We observed that NonSevere group patients had less tachypnoea compared to Severe & Survived and NonSurvived group of patients. It suggested that, respiratory rate on admission is one of the prognostic indicators in severity of illness.^[20]^

### Laboratory markers and Clinical Severity in COVID-19 Pneumonia

Haemoglobin, Platelet count & total white blood cell count were not much different in all the categories on admission in current study population. In several studies higher Leukocyte count was found in critically ill patients. Lymphopenia and eosinopenia were observed in majority of cases.^[21]^ The Neutrophil-Lymphocyte Ratio (NLR) on admission was found statistically different among these four groups (*P*=0.001). The ratio was higher among the severe categories of cases.^[22]^

Inflammatory markers were found to be increased in severe category of COIVD-19 cases, which usually raised after day three of admission. Which is correlated with other published studies.^[23]^ As per Ji P et al^[23]^trend of elevated inflammatory markers were associated with early identification, prediction of disease severity and adverse outcome in the form of death. This study has analysed CRP, Ferritin, D-Dimer & LDH with Severity of the disease and adverse outcome. This study observed strong C-index (AUC) for LDH post admission with severity and death.^[24]^Among all inflammatory markers, LDH values after admission helped a lot in predicting severity.^[25]^ As per present study, LDH on day one of admission over 372unit/L is approximately 70% Sensitive and Specific in predicting severity of covid-19 pneumonia. This coincides with the data observed across several published studies. Same observation for LDH on day one of admission was seen with death. LDH value above 468u/L was 77% sensitive and 84% specific for death. Several studies suggested LDH having relation with early in-hospitaldeath.^[26]^ Even D-Dimer on third day of admission and seventh day of admission are related with early in-hospitaldeath.^[27]^ D-Dimer on third day, more than 1500mcg/L is 87% sensitive and 65% specific for death, while value on seventh day, more than 3500mcg/L is 69% sensitive and 86% specific for early in-hospital death. These values correlated with Zhou F et al^28^and other studies.^[29,30]^

Deficiencies in certain micronutrients, in particular Vitamin-D deficiency was discussed a lot in determining the severity of the disease, but was not found significantly affecting the outcome in our study population. Several studies demonstrated its relation with severity but it seems to be affected by several confounders.^[31,32]^

Advanced age (more than 50 years), Ischemic Heart Disease, Raised troponin are associated with excess mortality. Several studies demonstrated and expressed issues with raised troponin not to be used solely as a diagnostic marker of myocardial infarction and heart failure in COVID-19 illness. This might be due to injury caused by the virus to cardiac tissue rich in ACE-2 receptors.^[33]^

We, attempted to predict severity and in-hospital early outcome by formulating a regression formula. These needs to be validated on larger study population before using for routine clinical utility.

## CONCLUSION

Advanced age, male gender, IHD, Respiratory Rate & Heart Rate on admission were associated with severe covid-19 illness. LDH & D-Dimer were associated with severe covid-19 illness and early in-hospital death.

## Supporting information

Institutional Review Board Approval Letter

## Data Availability

Data is available with hospital and statistical datasheet is available for sharing in case needed after approval from Institutional Review Board.

## ETHICS STATEMENT

This study was conducted independently without any external source of funding. Highest code of conduct was followed with utmost credibility ensured. The study was conducted after approval from Institutional Review Board (NHL Institutional review Board). Patient data were anonymized to maintain confidentiality of patient privacy.

## Acknowledgment

Authors would like to thank Dr. Pratik Patel, The Dean Smt NHL Municipal Medical College, Dr. Sundeep Malhan, The Medical Superintendent, SVPIMSR and Dr. Ami Parikh, HOD, Dept. of Medicine, Smt NHL Municipal Medical College for their constant guidance and support. We also acknowledge residents of our unit for their contribution in gathering data. We are thankful to our esteemed patients.

