## Supplementary material for "“Urban Tertiary Care Centre Experience of Characteristics of Severe COVID-19 Pneumonia”": Institutional Review Board Approval Letter

### NHL INSTITUTIONAL REVIEW BOARD (NHLIRB)


#### SMT N H L MUNICIPAL MEDICAL COLLEGE

Ellisbridge, Ahmedabad Gujarat (India): 380 006. Phone: 079-26576275, Fax: 079-26578451, Website: www.nhlmmc.edu.in



**Members:**

**Non Institutional**

Dr B D Mankad
Chairperson,
Prof & Head
Medicine, GCS

Dr Asit Patel
Member, Clinician
Prof of Surgery, AMCMET

Dr Nilay Suthar
Member, Clinician
Prof of Medicine, AMCMET

Mr Narendra Shah
Member Lay person
Social Worker

**Institutional**

Dr Pratik Patel
I/c Institutional Head &
Dean NHLMMC

Dr Deepa Banker
Prof of Pediatrics
SVP

Dr RajalThaker
Prof of Obs&Gyn, SCLGH

Dr Nadim Lil
Prof of Orthopedics,
SVP

Dr Nilima Shah
Assoc. Prof.,
Dept.of Psychiatry
SVP, NHLMMC

Dr Shashwat Jani
Assoc. Prof
Dept of ObGy
NHLMMC

Dr Neeraj Mahajan
Ass Prof
Dept of Physiology
NHLMMC

Dr Devang Rana
Asst Prof
Dept of Pharmacology
NHLMMC

Dr Tanmay Mehta
Asst Prof
Dept of Microbiology
NHLMMC

Mr Amit Dave
Legal Advisor
AMC

**Member Secretary:**

Dr Falguni Majmudar
Member Secretary
Asst Prof
Dept of Pharmacology
NHLMMC


**Date:11/09/2020**

To
**Dr. Janak Khambholja,**
Professor,
Department of General Medicine,
SVPIMSR, NHLMMC

**Ref:" Comparative study of efficacy of various adjunct treatment options in adult COVID-19 illness: A Case Control Study "**

**Dear Dr. Janak Khambholja,**
At the meeting held on 9th **September, 2020**, the Institutional Review Board, Smt. NHL Municipal Medical College [NHLIRB] reviewed and discussed your application for approval of your research study titled, " **Comparative study of efficacy of various adjunct treatment options in adult COVID-19 illness: A Case Control Study"** along with your co investigators Dr. Nehal Shah and Dr. Nilay Suthar

The following documents were reviewed during the meeting:
***Study Protocol including case record form***

During the meeting, the members of the NHLIRB reviewed the submitted documents and discussed the study in detail. The investigators were able to answer the queries raised by the committee satisfactorily.
A sub study **"Characteristics of severity in COVID-19 Pneumonia: a retrospective Observational Study"** from the data collected there was also proposed by your study team.

The committee decided to collectively **approve** the conduct of both the above studies in the presented form.

The following members of the committee were present/voted in the meeting and were involved in the approval process:

**Dr B D Mankad, Chairperson, Prof & Head, Dept. of Medicine, GCS**
**Dr Nadim Lil, Prof of Orthopedics, SVP, NHLMMC**
**Dr Nilima Shah, Assoc. Prof., Dept.of Psychiatry, SVP, NHLMMC**
**Dr Shashwat Jani, Assoc. Prof., Dept of ObGy.,NHLMMC**
**Dr Falguni Majmudar, Member Secretary, Asst Prof., Dept of Pharmacology, NHLMMC**
**Dr Tanmay Mehta, Asst Prof., Dept of Microbiology, NHLMMC**
**Dr Devang Rana, Member, Asst. Prof., Dept of Pharmacology, NHLMMC**
**Mr Narendra Shah, Member, Social Worker**
**Mr Amit Dave, Legal Advisor, AMC**

**Please Note:**
***You are requested to update NHLIRB about the progress of the study. You are directed not to violate the protocol and inform NHLIRB about protocol deviation immediately. You are also directed to intimate NHLIRB any change/ amendment in Protocol/Informed consent/Questionnaire and submit FINAL REPORT of the study at the completion of the study.***

FMajmudar

**Dr Falguni Majmudar**
**Member Secretary**
**Date:** 11/09/2020


Scanned with CamScanner
